# Optimising scan body enhances accuracy of full-arch implant scan using a smartphone video with deep learning model: An in vitro study

**DOI:** 10.64898/2026.08.10.26360076

**Authors:** Yuqing Lu, Jiayi Yu, Fei Liu, Tim Joda, Junying Li

## Abstract

**Objective:** A deep learning (DL) model was used to convert smartphone videos of a complete arch implant cast into 3D scans. The aim of current study was to determine if a custom scan body (SB) with geometric features and coating would outperform regular PEEK stock SB in this DL scenario. The DL-derived scan outcomes were compared with those obtained from a conventional splinted open-tray impression and from photogrammetry.

**Materials and Methods:** A maxillary edentulous model with six implants and multi-unit abutment analogs was scanned using four protocols: conventional splinted open-tray impression (CO), photogrammetry (PG; Icam4D), DL using stock SBs (DLS) and DL using custom SBs (DLC). Each protocol was repeated for 10 times. The DL scans were produced from smartphone videos with a high-fidelity, multi-view 3D construction AI model (Neuralangelo). The custom designed SB incorporated geometric features and was fabricated via 3D printing followed by a spray coating. Accuracy (trueness and precision) was assessed using three measurements: Root Mean Square (RMS), linear deviation, and angular deviation.

**Results:** DLC outperformed DLS in both trueness and precision regarding RMS and linear measurements (p<0.001). CO and PG demonstrated the highest RMS and linear trueness, with no significant difference between them (RMS: p=0.93; linear: p=0.663). PG achieved the best precision across RMS, linear and angular measurements.

**Conclusion:** The optimised SB significantly improves the accuracy of DL-based approach for full-arch implant scan comparing to regular PEEK stock scan bodies. While early stage, neural surface reconstruction has potential as a viable option for full-arch implant rehabilitation.

## 1. Introduction

In full-arch implant rehabilitation, obtaining an accurate impression is a critical factor for ensuring a passive fit of the prosthesis and long-term success of the treatment (Flügge et al. 2018; Abdelrehim et al. 2024). The current gold standard of conventional splinted open-tray impression followed by desktop scanning, is both time-consuming and technique-sensitive (Lyu et al. 2025a; Abuduwaili et al. 2025; Cheng et al. 2024; Gómez-Polo et al. 2023a), prompting interest in direct digital alternatives. Intraoral scanners (IOS) represent a potential alternative; however, their use as a sole modality for full-arch implant impressions remains controversial due to cumulative stitching errors over long spans (Rutkūnas et al. 2025; Chen et al. 2025; Li et al. 2024; Liu et al. 2024). Photogrammetry offers a more precise digital approach by determining implant coordinates from extraoral recordings of optical markers on scan bodies (SBs) (Cheng et al. 2024; Lyu et al. 2025b; Schmidt et al. 2022; Ma et al. 2021). However, evidence regarding whether photogrammetry performs comparably to the conventional technique remains inconsistent (Gómez-Polo et al. 2023a). Furthermore, despite the high cost of photogrammetry systems, their inability to capture adjacent teeth and peri-implant soft tissues necessitates additional intraoral scanning and subsequent alignment (Revilla-León et al. 2025a). This situation underscores the demand for novel digital approaches that offer comparable accuracy with greater efficiency and accessibility.

With the rapid advancement of new techniques involving neural network and deep learning (DL), new possibilities for high-fidelity 3D scanning have been unlocked. A notable milestone in this field is Neural Radiance Fields (NeRF) introduced in 2021 by Mildenhall et al. (Mildenhall et al. 2021), which achieves photorealistic view synthesis through coordinate-based multi-layer perceptrons (MLPs). Among the DL approaches building upon this approach, Neuralangelo, proposed recently by Li et al. (Li et al. 2023), represents a breakthrough in dense 3D reconstruction from multi-view images. It samples 3D points along camera view directions, encodes their spatial positions using a multi-resolution hash-grid encoding. These features are passed into an SDF MLP and a color MLP, and the surface is synthesised via an SDF-based volume rendering. Such network undergoes iterative training to refine surface geometry progressively, leading to substantial improvements in the quality of reconstructed surface meshes (Li et al. 2023; Yao et al. 2024; Farhat et al. 2025).

The potential of DL-based, multi-view 3D construction for dental scanning remains largely unexplored. A preliminary investigation by our research team represented the first attempt to apply the Neuralangelo model to full-arch implant scanning (Li et al. 2025). In that study, a smartphone was used to record a maxillary model with six implants, and the resulting datasets were used to train the DL model. The reconstructed implant model showed trueness comparable to that of an IOS. However, the limited reliability of IOS for full-arch implant impressions indicates the need for further improvement. It was also found that artifacts appeared on the homogeneous surfaces of the stock SBs rather than on the model itself. This phenomenon is considered an inherent limitation of the DL model, which has difficulty reconstructing regions with low surface texture or uniform coloration (Li et al. 2023). These artifacts or inaccurate scan may lead to misalignment of the SB and consequently inaccurate implant positions (Vandeweghe et al. 2017). Therefore, the use of optimised SBs with enhanced surface texture is expected to mitigate DL-generated artifacts and improve scanning accuracy of implant positions.

In this study, a custom SB was designed and fabricated tailored for the DL-based scanning technique that combines a neural surface construction model and smartphone videos, aiming to further explore its potential in scanning accuracy for full-arch implant rehabilitation. The results were compared to the scan generated by the same technique with a stock SB, as well as to that obtained via photogrammetry and conventional technique. The baseline reference was established through a direct scan of the master model using a desktop scanner.

## 2. Materials and Methods

This study has followed The Strengthening The Reporting of Observational Studies in Epidemiology (STROBE) Statement (Appendix 1). This is an in vitro study so ethics approval was not required. This in-vitro study was conducted on a model without human subjects or specimens; therefore, ethics approval was not required. As illustrated in Figure 1, an edentulous maxillary stone cast with six multi-unit abutment analogs (Analog for screw-retained abutment, Ø 4.6mm; Institute Straumann AG, Basel, Switzerland) was used as the master model. The implants were placed in the edentulous ridge at the regions of two lateral incisors, two first premolars, and two first molars. The model was scanned using for different protocols: conventional open-tray impression with splinting framework (CO), photogrammetry (PG), a DL-based approach with stock SBs (DLS), and the DL-based approach with custom SBs (DLC). Ten repeated scans were obtained per group (n=10). The model was also digitised using a desktop scanner (LS3; Kavo, Biberach, Germany) as the reference.

**Figure 1.**
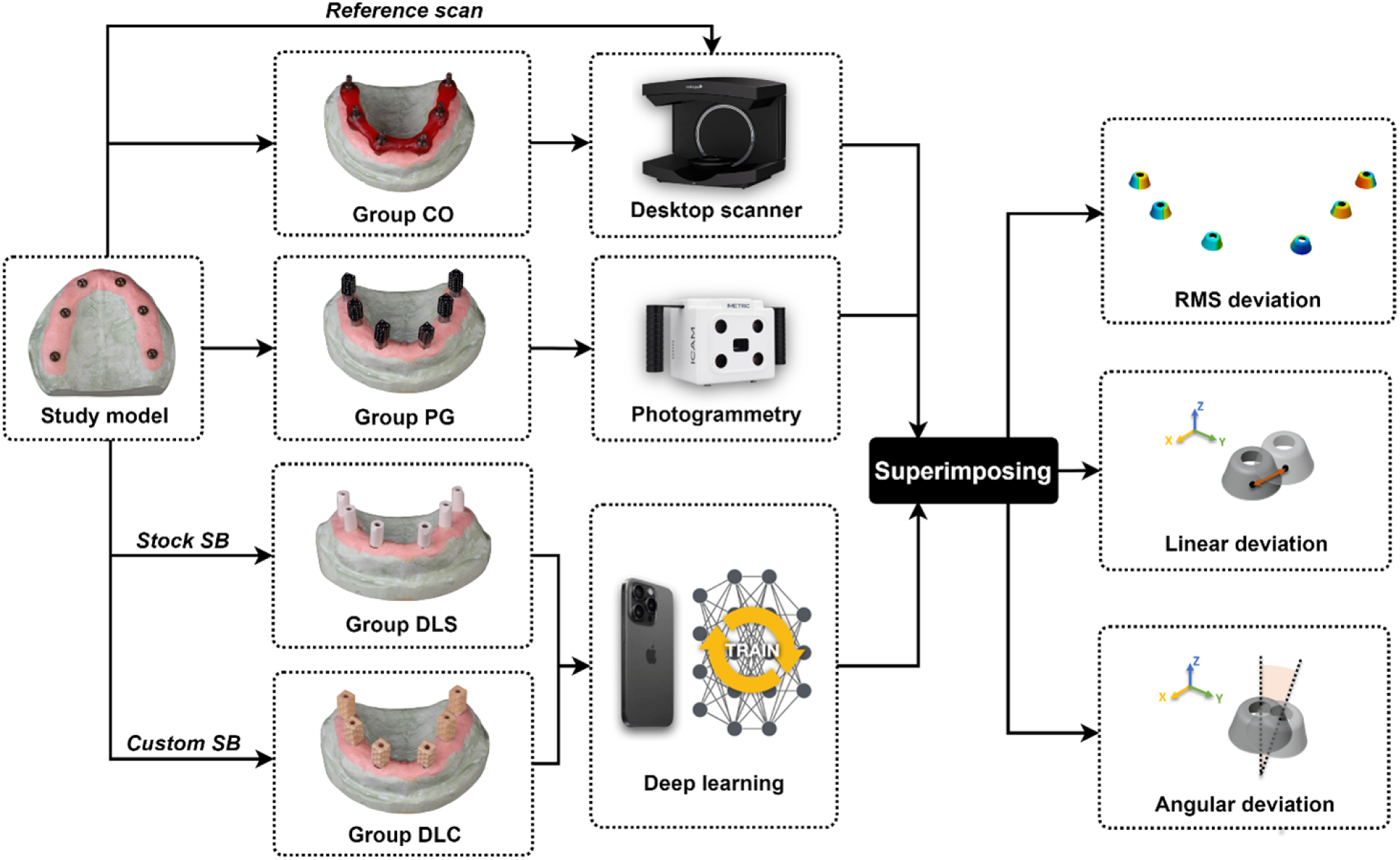
Flowchart of the study design. The model was scanned using four protocols: a conventional open-tray impression with a splinting framework (CO), photogrammetry (PG), a deep learning-based approach with stock SBs (DLS), and a deep learning -based approach with custom SBs (DLC).

For group CO, six open tray impression copings were on the multi-unit abutment tightened at 10 Ncm. Then they were splinted together with dental floss and self-consolidating acrylic resin (Unifast Trad; GC Corporation, Tokyo, Japan). The splinted framework was sectioned between each implant and linked again using a minor amount of same resin to minimize tension caused by shrinkage. The conventional impression was taken using a double-mix technique with polyvinyl siloxane impression material (Aquasil Ultra Medium and XLV; Dentsply Sirona, York, PA, USA). Multiunit abutment analogs were attached to the copings, and the master cast was made by pouring the impression with type IV dental stone. Six stock SBs (CARES Mono Scanbody; Institute Straumann AG, Basel, Switzerland) made from polyether-ether-ketone (PEEK) were screwed onto the analogs with a torque of 10 Ncm. The cast was digitised using the desktop scanner.

Regarding group PG, a photogrammetry system (Icam4D Camera, Generation 4; Imetric, Courgenay, Switzerland) was used. Before operation, a 30-minute warm-up process was completed according to the manufacturer’s instruction. Six SBs with optical markers (ICamBody, Imetric4D; Imetric, Courgenay, Switzerland) were placed on the multiunit abutment analogs of the model, with edge between two faces oriented anteriorly, and tightened at 10 Ncm. Before each scan, the SBs were calibrated using a calibration plate. Scans were captured by using the capturing camera with a scanning distance of approximately 30 cm (Revilla-León et al. 2024). Scanning was completed once all SBs indicated a green status in the software, resulting in about 40 views. All the scanning data were subsequently exported in STL format for further processing.

In terms of DL-based groups, two types of SBs were used respectively: stock PEEK SBs (Group DLS) and custom SBs designed and fabricated in this work (Group DLC). For the custom SBs, the upper part contained geometries were created using an industrial CAD software (Blender), while the bottom and screw channel was designed in a dental CAD software (DentalCAD; exocad) (Figure 2). The designed SBs were exported in STL. File. After, a dental polymer 3D printer (SprintRay Pro 95; SprintRay, CA, USA) was used to fabricate the SBs using a model material (SprintRay Die and Model tan 2; SprintRay, CA, USA). Supports were created on the top of the SB to ensure the bottom fit and maintain its structural integrity. The layer thickness was set to 50 μm. The printed SBs were cleaned in 91% Isopropyl in a washing machine (ProWash; SprintRay, CA, USA) followed by light curing (ProCure 2; SprintRay, CA, USA) and support removal. The fabricated SBs were visually checked for structural integrity, surface details and fitness on the multi-unit abutments. The qualified SBs were selected and coated with a matte spray coating with to eliminate the effect of material translucency on scanning accuracy.

**Figure 2.**
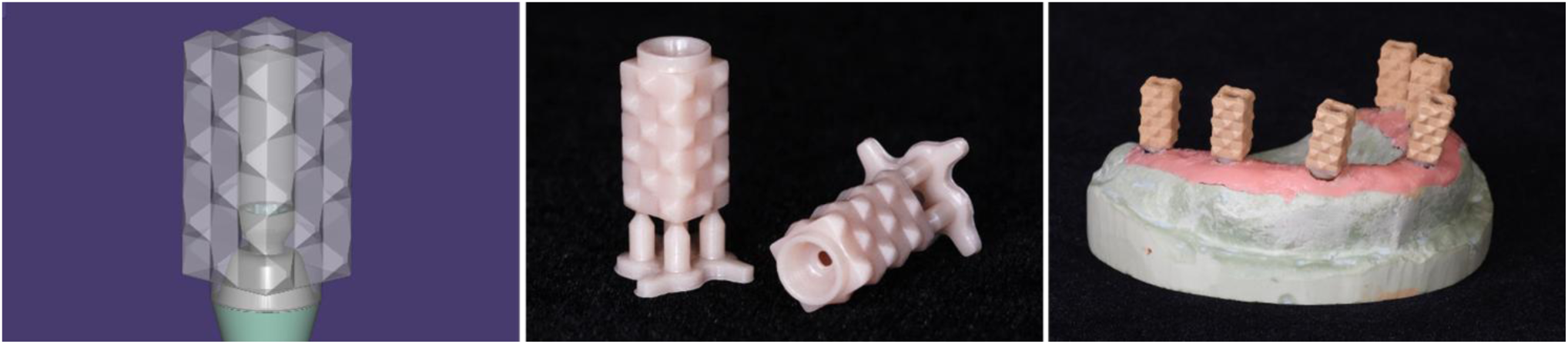
Digital design and fabrication of the custom SB. (a) 3D model of the multi-unit level custom SB. (b) polymer custom SBs fabricated via digital light processing. (c) Fully-seated custom SBs on the master model after spray coating.

The repeatability of the custom SBs was assessed in a pilot study. Six custom SBs and six stock SBs were placed individually on another cast model with only one multi-unit analog and was scanned using a desktop scanner. All the scans were superimposed using the model area while excluding the SB area. For each type of SB, the distance between the SB bottom central points of every two SBs were calculated and analysed using statistical software (IBM SPSS Statistic, v26.0; IBM Corporation, Armonk, NY, USA). No statistical difference was found between the two types of SBs (Figure 3), indicating a comparable repeatability of the custom SBs to the stock SBs.

**Figure 3.**
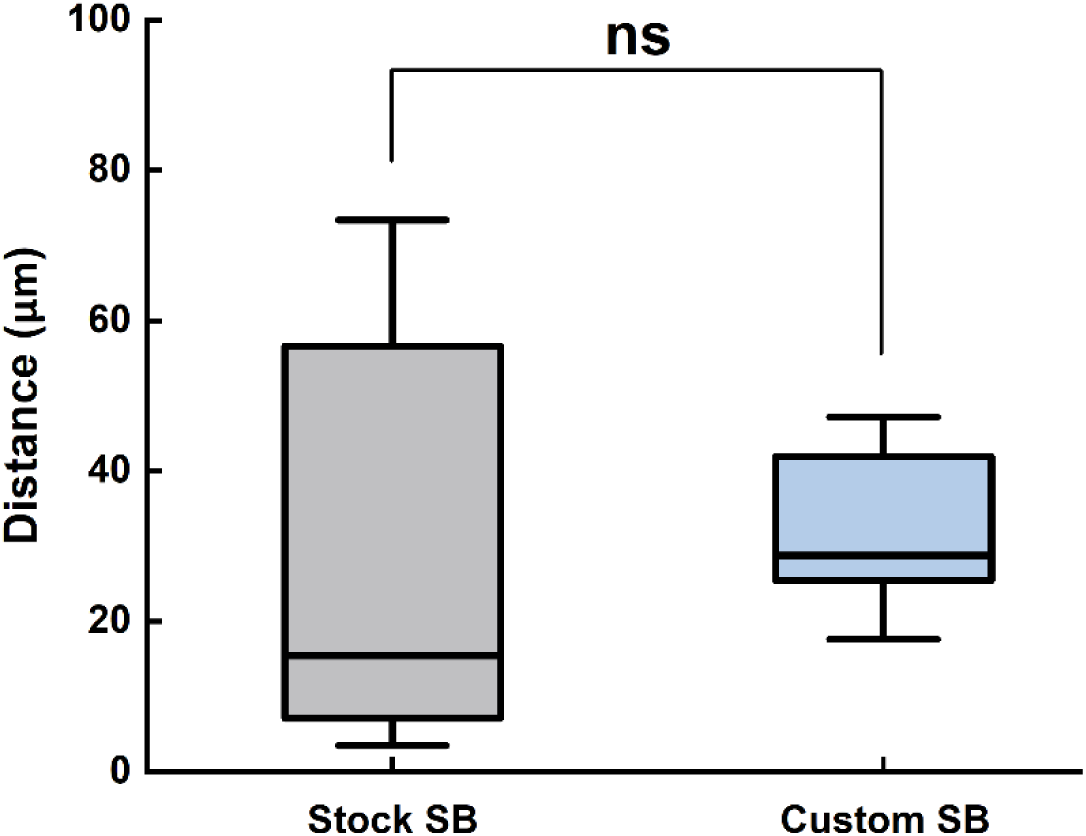
Repeatability analysis indicated no significant difference between the stock and custom scan bodies (SBs). The whiskers extend to the minimum and maximum values measured.

The verified six SBs of each type were placed on the implant model and hand-tightened. Then a smartphone (iPhone 15 Pro Max; Apple, CA, USA) with a 12-megapixel ultrawide camera was used to capture a video around the model. During recording, the mobile phone was held approximately 10 cm from the model and rotated 360 degrees in four perspectives: from left to right and from front to back. Each video was recorded at 4K resolution (24 frames per second) with a duration of approximately 40s and saved in MOV format, which were subsequently transcoded to MP4 format with a resolution of 1440p.

The DL-based approach generated 3D models in three stages (Figure 4) including dataset preparation, DL training and mesh extraction, as described in the previous study (Li et al. 2025). The videos were processed using the open-source Python software COLMAP to produce a dataset suitable for neural network training. A down-sampling rate of 2 was applied, and the scene type was set to object, yielding approximately 500 images. Subsequent analysis was performed in Jupyter Notebook, which enabled visualization of the camera poses, the reconstructed model and a spherical (Figure 4). The spherical boundary referred to the defined area for DL training, and its XYZ coordinates and overall scene scale were adjusted as needed to ensure that the model was centered within this region. A high-fidelity neural surface reconstruction model (Neuralangelo) was trained using the generated datasets with the following hyperparameters: batch_size = 2, dict_size = 22, and dim = 8. Model checkpoints were saved at 3000 training epochs, from which a dense 3D mesh was extracted at a resolution of 900 with a block size of 128 and subsequently converted to STL format. For each type of SB, 10 videos were recorded, leading to 10 extracted meshes as shown in Figure 5.

**Figure 4.**
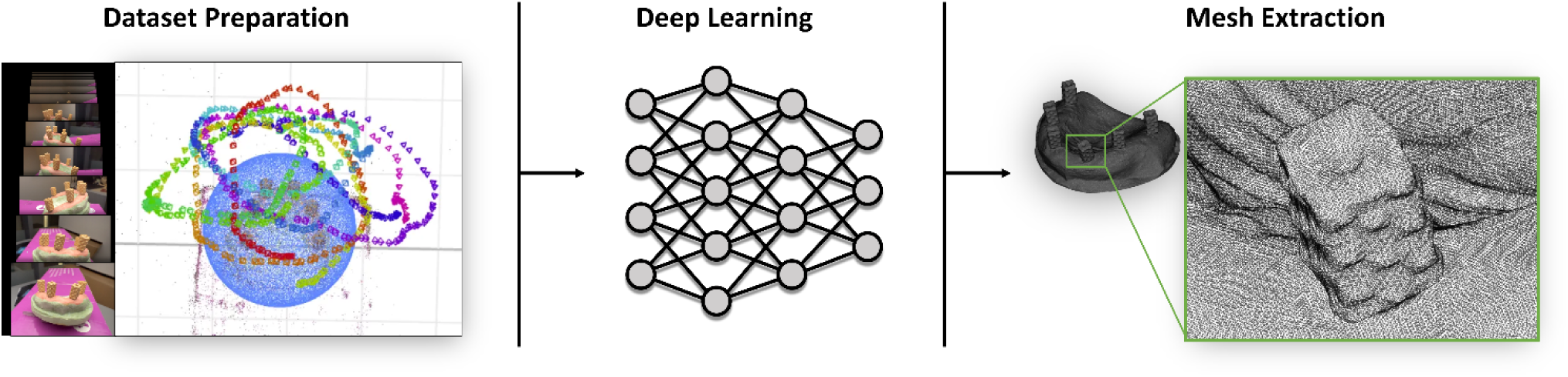
Workflow of the deep-learning based approach for obtaining full-arch implant impression from smartphone videos.

**Figure 5.**
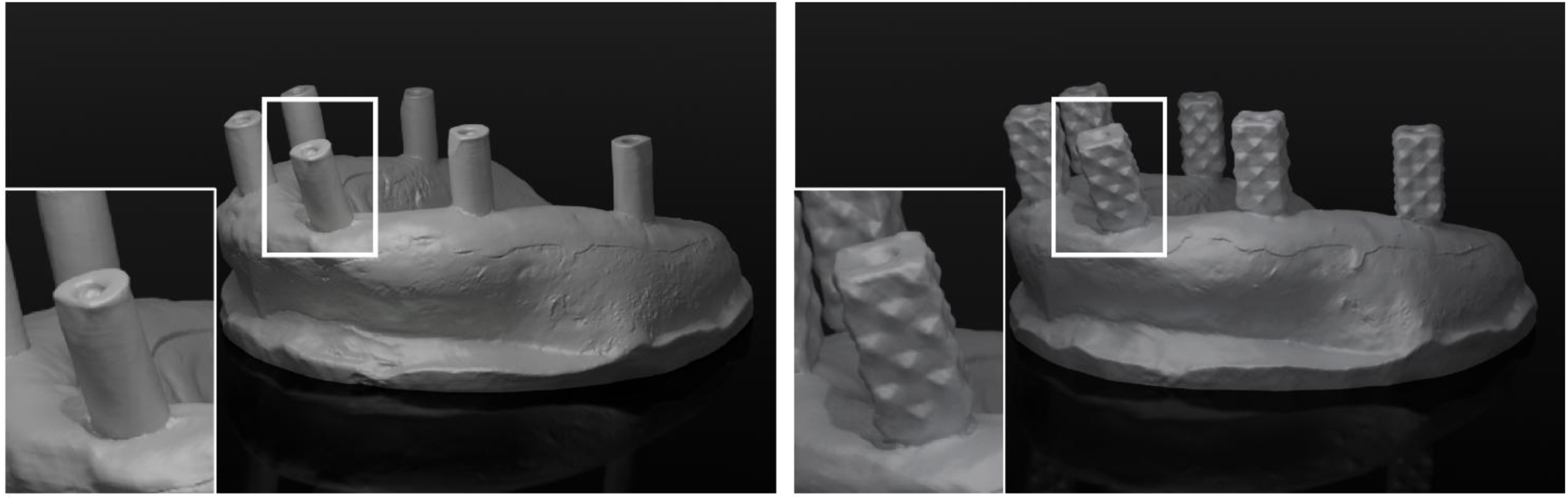
Scans obtained by deep-learning based scanning approach using stock SBs (a) and custom SBs (b). While artifacts were visible on both scans, the custom SB avoided large uniformly coloured areas, thereby reducing the risk of large distortion.

A total of 40 digital impressions were obtained from conventional, photogrammetry and DL-based approaches. An SB alignment procedure was performed in exocad, following the standard step in the digital workflow used by the CAD software to determine the position of each multi-unit abutment/implant. Before alignment, all model and reference scans were cropped to remove oral tissues to eliminate the influence of irrelevant areas. Each scanned SB was then aligned with a corresponding standard STL file using a best-fit algorithm. The standard SB files for the DLS and CO groups were obtained from the exocad library. For the custom SBs, the standard file was generated from the prototype STL file used for SB fabrication (Figure 2).

Before accuracy measurement, different SB geometries were standardised by replacing the SB with the bottom surface of the SB exported from exocad. The accuracy evaluation consisted of both trueness and precision: the former indicates the discrepancy between test scan and reference, while the latter is defined as the deviation between the test scans. All the digital impressions were superimposed on the reference data by the best-fit alignment. Discrepancy was quantified in three approaches: Root Mean Square (RMS), linear and angular deviations. The RMS discrepancy was calculated using a 3D inspection software (Geomagic Control X; 3D Systems, Morrisville, NC, US). For linear and angular measurement, a custom Python script was developed to quantify the deviations. The linear distance between SBs was determined based on the coordinates of their base central points, while angular deviation was calculated from the axes defined by the central points at the top and base.

The sample size was verified using G*Power to ensure a statistical power greater than 80%. Accuracy and precision data obtained from different measuring approaches were first tested for normality using the Kolmogorov–Smirnov and Shapiro–Wilk tests with SPSS software. For RMS Trueness, Welch’s analysis of variance (ANOVA) was used to compare the difference between the scanning protocols due to the presence of inhomogeneous variances. For RMS precision, Kruskal–Wallis test was adopted due to the presence of non-normality. For linear and angular measurements, a linear mixed model was utilised to assess trueness and precision. Post hoc comparison was subsequently performed for each measurement method. The significance level was set at α = 0.05.

## 3. Results

A total of 40 scans and 240 implant positions from the four evaluated scanning protocols were analysed. Table 1 and Figure 6 summarize the results obtained by RMS, linear and angular measurements. Significant differences were found between the scanning protocols for all the measurements (RMS trueness: p<0.001; RMS precision: p<0.001; linear trueness: p<0.001; linear precision: p<0.001; angular trueness: p<0.001; angular precision: p<0.001).

**Figure 6.**
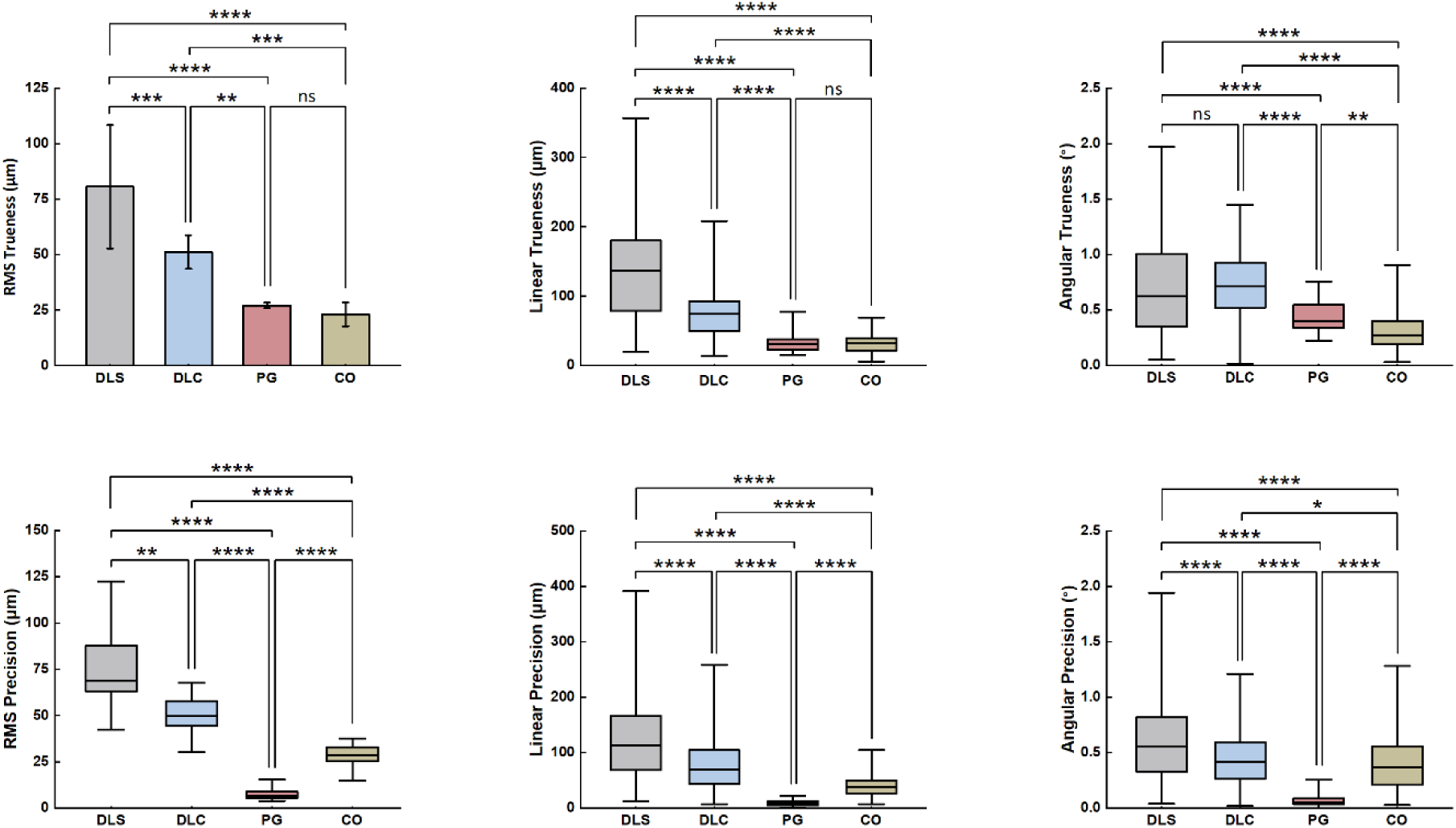
Trueness and precision regarding Root Mean Square (RMS), linear and angular deviations. The whiskers extend to the minimum and maximum values measured.

**Table 1.** Discrepancies of the SB positions obtained from the four evaluated scanning protocols.

| Deviation | Accuracy | n | Group | Mean | SD | Median | Min | Max | IRQ |
| --- | --- | --- | --- | --- | --- | --- | --- | --- | --- |
| RMS (μm) | Trueness | 10 | DLS<sup>a</sup> | 80.6 | 27.8 | 77.2 | 28.6 | 118.6 | 42.8 |
|  |  |  | DLC<sup>b</sup> | 51.1 | 7.6 | 49.3 | 40.1 | 66.9 | 10.0 |
|  |  |  | PG<sup>c</sup> | 27.1 | 1.3 | 26.9 | 25.0 | 29.1 | 2.4 |
|  |  |  | CO<sup>c</sup> | 23.1 | 5.4 | 21.8 | 16.7 | 31.9 | 10.7 |
|  | Precision | 45 | DLS<sup>a</sup> | 75.3 | 19.7 | 69.1 | 42.2 | 122.4 | 26.3 |
|  |  |  | DLC<sup>b</sup> | 50.5 | 9.6 | 49.8 | 30.0 | 67.8 | 13.8 |
|  |  |  | PG<sup>d</sup> | 7.9 | 3.3 | 6.7 | 3.9 | 15.4 | 3.4 |
|  |  |  | CO<sup>c</sup> | 28.4 | 5.3 | 28.5 | 14.9 | 37.7 | 7.5 |
| Linear (μm) | Trueness | 60 | DLS<sup>a</sup> | 144.1 | 83.4 | 136.5 | 19.5 | 356.8 | 106.1 |
|  |  |  | DLC<sup>b</sup> | 78.0 | 43.5 | 74.8 | 13.5 | 208.5 | 45.3 |
|  |  |  | PG<sup>c</sup> | 35.7 | 31.4 | 31.4 | 15.1 | 77.4 | 15.8 |
|  |  |  | CO<sup>c</sup> | 32.4 | 14.1 | 32.0 | 5.8 | 69.4 | 17.9 |
|  | Precision | 270 | DLS<sup>a</sup> | 122.8 | 71.3 | 112.9 | 12.5 | 391.5 | 97.4 |
|  |  |  | DLC<sup>b</sup> | 78.9 | 47.5 | 69.4 | 6.9 | 258.7 | 62.0 |
|  |  |  | PG<sup>d</sup> | 8.7 | 4.2 | 8.5 | 0.5 | 22.0 | 6.6 |
|  |  |  | CO<sup>c</sup> | 39.7 | 18.6 | 37.7 | 7.1 | 104.8 | 23.8 |
| Angular (°) | Trueness | 60 | DLS<sup>a</sup> | 0.71 | 0.44 | 0.62 | 0.05 | 1.97 | 0.67 |
|  |  |  | DLC<sup>a</sup> | 0.71 | 0.31 | 0.71 | 0.02 | 1.44 | 0.41 |
|  |  |  | PG<sup>b</sup> | 0.45 | 0.16 | 0.40 | 0.22 | 0.76 | 0.22 |
|  |  |  | CO<sup>c</sup> | 0.32 | 0.20 | 0.27 | 0.03 | 0.91 | 0.22 |
|  | Precision | 270 | DLS<sup>a</sup> | 0.61 | 0.36 | 0.56 | 0.04 | 1.94 | 0.49 |
|  |  |  | DLC<sup>b</sup> | 0.45 | 0.25 | 0.42 | 0.02 | 1.21 | 0.33 |
|  |  |  | PG<sup>d</sup> | 0.07 | 0.05 | 0.05 | 0.00 | 0.26 | 0.05 |
|  |  |  | CO<sup>c</sup> | 0.41 | 0.26 | 0.37 | 0.03 | 1.28 | 0.35 |

Note: Different superscript letters within the same column and same measurement denote statistically significant differences (p<0.05).
\*DLS: Deep learning using stock SBs. DLC: Deep learning using custom SBs. PG: Photogrammetry. CO: Conventional splinted open-tray technique. SD: Standard deviation. Min: Minimum. Max: Maximum. IRQ: Interquartile range.

Regarding RMS trueness, DLS group exhibited the greatest mean deviation of 80.6 ± 27.8 µm, whereas DLC showed a significantly improved trueness of 51.1 ± 7.6 µm (p<0.001); PG and CO groups demonstrated the lowest mean deviations (27.1 ± 1.3 µm and 23.1 ± 5.4 µm, respectively), with no statistically significant difference between the two groups (p=0.93). For RMS precision, DLS group again showed the highest discrepancy, with a median deviation of 69.1 µm and a range of 42.2–122.4 µm, followed by DLC (median: 49.8 µm, range: 30.0–67.8 µm, p=0.001); the lowest discrepancy was observed for PG, with a median of 6.7 µm and a range of 3.9–15.4 µm, followed by CO (median: 28.5 µm, range: 14.9–37.7 µm, p<0.001).

Linear deviations demonstrated consistent result as RMS discrepancy. In terms of trueness, DLS group showed the largest median deviation of 136.5 µm, ranging from 19.5 to 356.8 µm, whereas DLC presented a significantly lower deviation (median: 74.8 µm, range: 13.5–208.5 µm, p<0.001); PG and CO groups produced the smallest deviations, with median values of 31.4 µm and 32.0 µm, and ranges of 15.1–77.4 µm and 5.8–69.4 µm, with no significant difference (p=0.663). Regarding linear precision, DLS remained the least precise, presenting a median deviation of 112.9 µm and a range of 12.5–391.5 µm, while DLC showed a reduced discrepancy (median: 69.4 µm, range: 6.9–258.7 µm, p<0.001); PG group achieved the best precision with a median deviation of 8.5 µm and a range of 0.5–22.0 µm, followed by CO (median: 37.7 µm, range: 7.1–104.8 µm, p<0.001).

In terms of angular deviations, CO reached the highest trueness (median: 0.27°, range: 0.03°–0.91°), followed by PG (median: 0.40°, range: 0.05°–1.97°, p=0.004); in contrast, DLS and DLC showed similarly higher median deviations of 0.62° (range: 0.05°–1.97°) and 0.71° (range: 0.02°–1.44°) (p=0.945). With respect to angular precision, PG demonstrated the lowest deviation (median: 0.05°, range: 0.00°–0.26°), whereas DLS exhibited the greatest discrepancy (median: 0.56°, range: 0.04°–1.94°); CO group showed slightly lower median deviation (median: 0.37°, range: 0.03°–1.28°) than DLC (median: 0.42°, range: 0.02°–1.21°, p=0.032).

## 4. Discussion

This study sought to enhance the scanning accuracy of full-arch implant impression generated by a deep learning model and a smartphone-captured video through the optimisation of SB design. A custom SB was designed and fabricated, resulting in significantly improved accuracy in both trueness and precision based on RMS and linear measurements comparing with the regular PEEK stock SB. Although the achieved accuracy remains inferior to that of the established conventional technique and photogrammetry, these findings highlight the potential of DL-based reconstruction method as a digital impression technique and the need for further refinement to support its clinical application in full-arch implant rehabilitation.

The geometry of scan bodies plays a notable role in the accuracy of scanned implant positions (Gómez-Polo et al. 2023b; Pan et al. 2025; Michelinakis et al. 2024; Lawand et al. 2024). However, no clear agreement has been reached regarding the optimal SB design. One study using a laboratory scanner showed that SBs with a length above 8 mm and a diameter above 4.8 mm are more accurate than smaller sizes (Pan et al. 2025). It also indicated a higher linear accuracy for the cylindrical SB and a higher angular accuracy for the cuboidal design. Regarding ISO, a study demonstrated that less complex SB geometries are associated with higher congruence and reduced stitching error (Michelinakis et al. 2024), while other studies suggested that the scanning accuracy can be improved by adding geometric features on SB (Lee et al. 2021; Lawand et al. 2024). In the present study, the stock SB had a diameter of 4.6 mm and a height of 10 mm, while the custom SB had a dimension of 5 mm x 5 mm x 11 mm. The results demonstrated that, the custom SB with cuboidal shape, increased dimensions with additional geometric features that led to significant improvement in RMS and linear accuracy. Further, there was a moderate improvement in angular precision. This may be attributed to more stable feature detection during DL-based reconstruction, reducing the influence of surface artifacts and facilitating more reliable alignment of the SBs.

Beyond geometry, the material properties of SBs also affect scanning accuracy. Previous studies have shown that optical characteristics such as surface reflectiveness and translucency can influence the accuracy for dental scanners (Li et al. 2017; Arcuri et al. 2020; Lee et al. 2021; Chen et al. 2025). Accordingly, an opaque matte spray coating was utilized to minimize the effects of translucency of the 3D-printed polymer. Regarding mechanical properties, materials with lower elasticity are less prone to deformation during tightening, leading to more consistent seating and better positional repeatability (Arcuri et al. 2020; Gómez-Polo et al. 2023b). Despite being fabricated from a different material, the custom and PEEK SBs in this study demonstrated similar repeatability. However, it should be noted that the adopted polymer material, as a dental model resin, has a low resistance to fatigue and aging compared with commonly used SB materials such as PEEK and titanium.. Fabrication of the custom SB design in these more durable materials should therefore be considered for repeated clinical use.

Although the accuracy of the DL-based approach was significantly improved, it remained inferior to both the conventional technique and the photogrammetry system evaluated in this study. The deviations measured for the conventional workflow fell within the expected range (trueness: 34–72 µm, precision: 9–73 µm) for this established gold standard (Ma et al. 2021; Liu et al. 2024; Lyu et al. 2025b; Abuduwaili et al. 2025). The photogrammetry results also aligned well with previously reported values for the same system, which demonstrated trueness ranging from 11–69 µm and precision ranging from 2–41 µm (Ma et al. 2021; Cheng et al. 2024; Revilla-León et al. 2025b; Abuduwaili et al. 2025). A systematic review further indicated that implant prostheses with vertical misfits up to 160 µm or horizontal misfits up to 150 µm do not lead to mechanical or biological complications (Abdelrehim et al. 2024). In the present study, the accuracy achieved by the DL-based approach approached the ranges observed for both conventional impressions and photogrammetry. This suggests that, once incorporated into a full production workflow, the DL-generated impression may have the potential in producing implant restorations with clinically acceptable fit. These findings highlight the promising potential of this novel technique, particularly when applied in cases involving a smaller number of implants.

Although both the DL-based approach and photogrammetry rely on extraoral multi-view images, they operate through fundamentally different mechanisms. Photogrammetry determines implant coordinates by capturing images of SB markers from multiple angles and reconstructing their 3D positions using calibrated geometric calculations; its accuracy depends mainly on camera calibration, image orientation, and the quality of detected target points (Rivara et al. 2016). In contrast, the deep learning approach requires a substantially larger number of images to generate a mesh of the SB geometry, after which the workflow proceeds similarly to IOS impressions. The scanning accuracy is influenced by factors such as video quality, the number of training iterations, and the alignment process (Li et al. 2025). Further research is needed to better characterize the variables affecting the performance of neural surface reconstruction methods.

Based on the results of this study, photogrammetry demonstrated reliable performance comparable to a conventional technique. In agreement with an earlier investigation (Lyu et al. 2025a), the RMS and linear trueness observed for photogrammetry was similar to that of the conventional technique. Furthermore, these findings showed higher RMS and linear precision than conventional techniques, which is also consistent with some previous reports (Ma et al. 2021; Cheng et al. 2024; Lyu et al. 2025a; Abuduwaili et al. 2025). This can be attributed that the reduced number of procedural steps in photogrammetric workflows limits the cumulative errors commonly associated with conventional impressions. Nevertheless, other researchers have reported greater overall discrepancies with photogrammetry compared with conventional technique (Revilla-León et al. 2021, 2023). It should be noted that the outcomes may be influenced by factors such as photogrammetry systems (Revilla-León et al. 2025b), scanning distances (Revilla-León et al. 2024), implant angulation and distribution (Liu et al. 2024), among others. Despite these variations, there is a growing body of evidence supporting photogrammetry as a valid and clinically acceptable digital alternative for full-arch implant rehabilitation (Ma et al. 2021; Gómez-Polo et al. 2023a; Cheng et al. 2024; Lyu et al. 2025b; Abuduwaili et al. 2025).

Among the three measurement approaches, RMS and linear deviation produced consistent trends when comparing the different scanning protocols. The RMS value is calculated as the square root of the mean of the squared distances between corresponding points on the scanned model and the reference surface, providing a measure of the overall magnitude of deviation between the two surfaces (Mehl et al.). In the present study, linear deviation was calculated as the difference between the central bottom points of each SB in the scan and reference models, whereas some studies calculated linear deviation based on cross-arch distances between pairs of scan bodies (Cheng et al. 2024; Liu et al. 2024). Although some investigations have reported similar trends when using RMS and linear deviations (Wu et al. 2024; Liu et al. 2024), discrepancies may occur (Fu et al. 2024). Angular deviation was also evaluated in this study, but the results did not correspond with the trend in linear or RMS deviations. Further research is needed to clarify the significance of different measurement approaches and their relationships to better interpret their clinical relevance and improve the comparability of findings across studies.

This study demonstrated the potential of an economical, DL-based approach for full-arch implant scan. Although its accuracy does not yet reach that of conventional impression or photogrammetry, the results show meaningful promise and indicate that further refinement could make this technique a viable alternative. A key limitation is that all scans were performed on a single model under controlled laboratory conditions; in clinical setting, factors such as limited access, inconsistent lighting, and saliva may influence scan quality and consequently the accuracy of implant position. Additionally, the adopted multi-view 3D reconstruction model requires substantial training time (Yao et al. 2024). Future work should therefore focus on improving neural surface reconstruction to enhance both accuracy and computational efficiency.

## 5. Conclusion

Optimizing SB significantly enhances the accuracy of a full-arch implant scan obtained by the neural surface reconstruction model and smartphone videos. Although its accuracy has not yet reached the level of conventional splinted open-tray impression or photogrammetry, these findings highlight the potential of this novel technique and indicate that further optimization may enable it a viable alternative. Photogrammetry demonstrated comparable trueness and superior precision in comparison with the conventional technique.

## Data Availability

All data produced in the present study are available upon reasonable request to the authors.

## Acknowledgements

This study was funded by American Academy of Implant Dentistry (AAID) Foundation Research Grant. The authors thank William Giannobile for critically reviewing the manuscript.

## Author contributions

Y.L. and J.L. conceived the ideas; Y.L., J.Y. and J.L. collected the data; Y.L., F.L., T.J. and J.L. analysed the data; Y.L. drafted the manuscript; and all authors critically reviewed the manuscript.

